# Chronic emotional stress and mediating role of Interleukine-6 in the association with cardiometabolic disorders in a multiethnic middle-aged and older US-population

**DOI:** 10.1101/2025.04.23.25326283

**Authors:** Asma Hallab, The Health and Aging Brain Study (HABS-HD) Study Team

## Abstract

**Introduction:** Chronic emotional stress is a well-recognized risk factor for psychiatric and cardiometabolic disorders. The mediating role of low-grade inflammation in older, ethnically diverse populations has never been studied.

**Methods:** The multiethnic ≥ 50-year-old study population is a subset of the Health and Aging Brain Study: Health Disparities (HABS-HD) study. Adjusted logistic and linear regression were used to assess associations. Statistical mediation analysis with non-parametric bootstrapping was used to determine the intermediate role of Interleukine-6 (IL-6).

**Results:** The study included 2,173 participants (50-92 years). Hispanic and Black participants disclosed higher chronic stress levels than White participants. Having a chronic stress total score ≥ six points is associated with 53% higher odds of disclosing concomitant cardiovascular disease (CVD) (adj.OR=1.53 [1.1-2.53]), 31% of Type-2 diabetes (T2DM) (adj.OR=1.31[1.06-1.62]), 23% of hypertension (adj.OR=1.23 [1.02-1.49]), and 30% obesity (adj.OR=1.3[1.09-1.55]). These associations were statistically mediated by IL-6 (12% (*p-value*_FDR_=0.012) of the association with CVD, 17% T2DM (*p-value*_FDR_<0.001), 18% hypertension (*p-value*_FDR_<0.001), and 29% obesity (*p-value*_FDR_=0.005)).

**Conclusions:** The study highlights a further aspect of the pathophysiological mechanisms involved in brain-body communication. While IL-6 partially explains statistical associations between chronic emotional stress and major cardiometabolic disorders, potential causal effects need to be explored in larger longitudinal studies.

## 1. Introduction

Chronic emotional stress, first described by Hans Selye in 1956, is a long-lasting psychological distress that might be triggered by intrinsic or extrinsic factors destabilizing a state of homeostasis. (1) It is to be distinguished from the acute stress response, also known as the fight-or-flight reaction, described by Walter Cannon in 1915. (2) While the acute stress reaction has a central evolutionary function ensuring survival and safety, (2) its prolongation over time into a chronic form presents a pathological state associated with higher risks of health adversities. (1) Chronic emotional stress induces a cascade of physiological reactions, including hormonal disturbance, (3) neurovascular and systemic dysregulations, (4) and neurobiological impairment, (5, 6) a significant risk factor for mental health disorders such as depression and anxiety. Depression is a major mood disorder widely prevalent, showing a rapid increase in its incidence and associates with higher healthcare and economic burdens worldwide. (7) Similarly, anxiety is a stress-associated psychological pathology highly prevalent as a single or comorbid disorder. (8, 9) Depression and anxiety are at the top of mental health disorders worldwide and affect different populations of all age groups. (7, 10) They share several stress-related biosocial risk factors, including childhood and lifetime adversities, (11) overwhelming workloads, (12) and natural or man-made disasters, (13, 14) in addition to a significant association with higher disability rates and mortality. (15)

Similar to mental health adversities, cardiovascular and metabolic disorders have shown a sharp increase in their incidence in the last decades, inducing an alarming rise in mortality rates worldwide. (15) The association between stress and cardiometabolic disorders is complex and bidirectional. While people with stress-associated psychiatric disorders are highly exposed to cardiometabolic risk factors, such as obesity, diabetes, and heart infarction, (16–18) the psychological and cardiovascular burdens of those pathologies play, on the other side, a significant causal role in the pathogenesis of the emotional state of stress, depression, and anxiety. (19, 20)

Several biological models have been explored to explain the pathophysiology of the interaction between psychological adversities and cardiometabolic risk factors. In addition to socioeconomic insecurities, behavioral patterns, hormonal dysregulations, and cerebrovascular pathologies, systemic (and neuro-) inflammation has been demonstrated to be significantly associated with the genesis of mental (21, 22) and cardiometabolic disorders. (23, 24) Our previous study showed significant interleukin-6-mediated associations between depression, anxiety, and major cardiometabolic disorders. Low-grade inflammation might, therefore, be a significant actor in the chronic stress-cardiometabolic disease axis, too. However, no study has yet explored this theory.

Persons at advanced age are highly exposed to psychological adversities such as depression, anxiety, and chronic stress owing to various etiologies and triggers. (25) Their vulnerability, exacerbated by increased frailty, social withdrawal, loneliness, and health issues, makes them a particular risk group. (25) Furthermore, people from disadvantaged ethnic backgrounds have a high propensity to experience stressful life events and are at higher risk of being exposed to neuropsychiatric and physical adversities. (26) Several epidemiological studies have reported higher odds of cardiometabolic risk factors in ethnic minorities and marginalized groups. (27) The limited opportunities to ensure convenient healthcare management (28, 29) make this group more exposed to chronic complications and subject to higher morbidity and mortality rates. (7)

The summation of all those unfavorable backgrounds makes studying chronic emotional stress in a multiethnic population of specific interest in order to better understand its association with cardiometabolic risk factors and the mediating role of inflammation in this particular group. Very limited studies explored the association between psychological distress and cardiometabolic risk factors in older adults. Furthermore, most reports tend to be restricted to dominant ethnic groups and younger populations, mainly in industrialized societies. There is also an urgent need to understand and mitigate biases related to underestimating ethnic disparities in the published literature. (30)

The main aim of this study was to fill this gap by exploring the association between chronic emotional stress and cardiometabolic risk factors in a multiethnic population of middle-aged and older adults and evaluating the mediating role of systemic inflammation, particularly IL-6, in this association.

## 2. Methods

The study was performed in compliance with the Strengthening the Reporting of Observational Studies in Epidemiology (STROBE) guidelines. (31)

### a. Study population

The study population is a subset of the Health and Aging Brain Study: Health Disparities (HABS-HD), a follow-up cohort of the Health and Aging Brain among Latino Elders (HABLE) study initiated in 2017 at the Institute for Translational Research (ITR) at the University of North Texas Health Science Center (UNTHSC), Fort Worth, Texas. Fifty years and older community-dwelling adults were recruited in community-related events, and the study has also been advertised in the media. Participants underwent clinical, neuropsychological, biological, and neuroimaging investigations at a 24- to 30-month interval. The initial aim of the NIH-funded study was to assess health disparities between Mexican Americans and non-Hispanic White Americans. (32) The inclusion of a further 1,000 Black Americans in 2021 allowed a broader assessment of the three largest ethnic groups living in the United States of America (White, Hispanic, and Black). Cases with type-1 diabetes, severe health conditions (cancer in the last 12 months), end-stage renal disease, chronic health failure, chronic obstructive pulmonary disease, serious mental illness (including alcohol and substance disorder), active infection, and dementia other than Alzheimer’s type were not eligible. All participants gave written informed consent. The current statistical analysis was performed between November 2024 and February 2025 and was, therefore, based on the 5^th^ data release of HABS-HD.

The study is restricted to baseline data and cross-sectional analysis owing to a high loss of follow-up and a lower incidence of cardiometabolic disorders at 24 months.

Procedures contributing to this work comply totally with the ethical standards of the relevant national and institutional committees on human experimentation and with the Helsinki Declaration of 1975, including the revision of 2013. Ethical approval was obtained from the local institutional review board (North Texas Health Science Center Institutional Review Board).

### b. Chronic emotional stress

Chronic emotional stress, dating over **<u>six months</u>**, is assessed using a self-administered questionnaire, which includes eight main questions (**Supplementary Table 1**). The **Chronic Stress Total score** was defined as the sum of Chronic Stress 1 through Chronic Stress 8c, except Chronic Stress 8a.

Depression and anxiety were included in the analysis as major predictors of psychological burdens and as intermediates in the path between chronic emotional stress and the outcome of interest. A participant was diagnosed with **depression:** if he/she reported “a past medical history of depression or relevant medication or his/her Geriatric Depression total Score (GDS) was ≥ 10 points”. **Anxiety** was diagnosed when the participant disclosed “a positive past medical history of diagnosed anxiety” or disclosed “relevant medication”.

### c. Cardiometabolic risk factors

A major cardiometabolic risk factor and surrogate biomarker of the following cardiometabolic diseases is **Body Mass Index (BMI)**, which is defined as the result of weight (Kg) / height (m)^2^. Glycated Hemoglobin A1c (HbA1c) was calculated from fasting blood samples and reported in percentages (%). Plasma levels of total cholesterol, high-density lipoprotein (HDL), and low-density lipoprotein (LDL) were measured in fasting blood and reported in mg/dL. Systolic and diastolic blood pressure measurements were performed at rest and reported in mmHg. The heart rate or pulse was also assessed at rest and reported in beats per minute (bpm).

Cardiometabolic diseases are reported as binary variables and defined in the study as follows:

- **Cardiovascular disease (CVD)**: is defined as “a positive past medical history of heart attack, heart failure, cardiomyopathy, atrial fibrillation, or heart valve replacement”.
- **Type 2 Diabetes Mellitus (T2DM)**: if “HbA1c ≥ 6.5 ***<u>OR</u>*** past medical history of diabetes, ***<u>OR</u>*** relevant medication”. Participants with type 1 diabetes were not eligible for the study.
- **Dyslipidemia** is defined as “Low-Density Lipoprotein (LDL) ≥ 120 mg/dL, ***<u>OR</u>*** total Cholesterol ≥ 240 mg/dL, OR Triglycerides (TG) ≥ 200 mg/dL, ***<u>OR</u>*** past medical history of high Cholesterol, ***<u>OR</u>*** relevant medication”.
- **Arterial hypertension** is defined as “Past medical history of hypertension, ***<u>OR</u>*** Consistent elevation of blood pressure across both measurements, ***<u>OR</u>*** at least two blood pressure readings of Systolic Blood Pressure (SBP) ≥ 140 mmHg or Diastolic Blood Pressure (DBP) ≥ 90 mmHg ***<u>OR</u>*** relevant medication”.
- **Obesity** was given as a binary diagnosis if the BMI was equal to or higher than 30.

### d. Systemic inflammation

Systemic inflammation was evaluated through IL-6 levels. IL-6 was measured in the fasting blood serum and reported in pg/mL. Measurement details were reported in a previous methodological publication. (32) Owing to their extreme deviation from the normal distribution (right skewness), IL-6 levels underwent a log transformation for the current analysis.

### e. Covariables

Age (years), sex (“female” vs. “male”), educational level (years), ethnicity (self-reported ethnicity of “non-Hispanic white”, “Hispanic”, or “Black”), Tobacco smoking (binary), alcohol consumption (binary), no physical activity (four cases with missing values were attributed to the group), and BMI are considered strong predictors of cardiometabolic disorders and were adjusted for in the regression models.

### f. Inclusion criteria

Only non-duplicated cases with complete data on chronic emotional stress, BMI, and IL-6 values were included (**Figure 1.a**).

**Figure 1:**
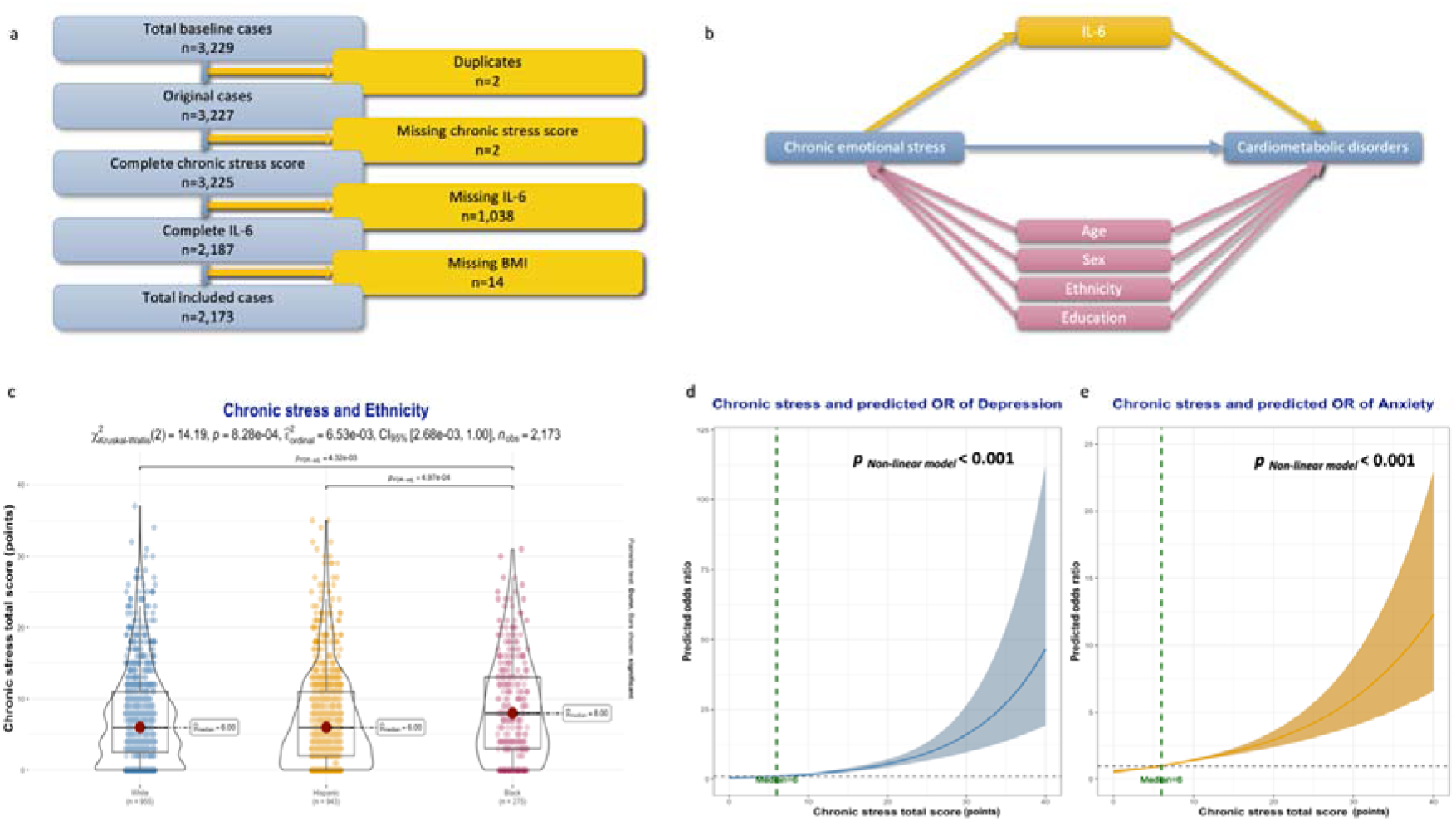
Inclusion criteria and characteristics of study participants. **1.a.** Flow chart of included cases. **1.b.** Directed acyclic graph detailing the variables included in the statistical mediation analysis. **1.c.** Differences between ethnic groups in chronic stress scores. **1.d.** Association between chronic stress levels and odds of depression. **1.e.** Association between chronic stress levels and odds of anxiety.

### g. Statistical analysis

The statistical analysis and data visualization were performed using RStudio version 2024.12.1. Continuous variables were reported in medians with interquartile ranges (IQR), and count variables were reported as numbers with percentages (%). Chronic stress-related group comparison was performed using the Wilcoxon rank sum test for continuous variables and Pearson’s Chi-squared (*X*^2^) test for count variables. *X*^2^ Kruskall-Wallis was applied to assess differences between ethnic groups regarding their chronic stress score levels, and the corresponding *p-values* were reported.

The association between chronic emotional stress and biological biomarkers of cardiometabolic risks was evaluated using linear regression models. When indicated, restricted cubic splines were included in the models to test for non-linearity.

The association between the exposure to chronic emotional stress as a binary independent variable (the median value as a cutoff) and concomitant cardiometabolic disorder as a binary dependent variable was assessed using logistic regression. The crude models were first reported with the corresponding odds ratios (*OR*), 95% confidence intervals (*CI*), and *p-value*. Adjusted models were then assessed using the same method and after including the predefined covariables.

Mediation analysis was performed using a 1000-fold non-parametric bootstrapping method of 95% CI, and the Average Causal Mediation Effects (ACME), Average Direct Effects (ADE), and total effect, as well as their corresponding 95% *CI* and *p-values*, were visualized. To simulate causal frameworks, statistical mediation models were adjusted only for covariables with confounding effects on the dependent and independent variables simultaneously (**Figure 1.b**). The percentage of mediated effect (rounded value without decimals) corresponded to the proportion of ACME from the Total Effect. The False Discovery Rate (FDR) method was used to reduce the risk of Type I errors. The resulting *p_FDR_*-values were reported. Two-sided *p*-and *p_FDR_*-*values* under 0.05 were considered statistically significant.

A sensitivity analysis was performed based on the total score of chronic emotional stress as a continuous independent variable.

## 3. Results

### a. Study population

The study included 2,173 participants aged between 50 and 92 years and with a median age of 66 (59, 72). Among them, 955 (44%) disclosed themselves as “white”, 943 (43%) as “Hispanic”, and 275 (13%) as “black”. Women represented 62% of the study population. The median value of the chronic stress total score was six points (2, 11).

Based on this median value as a cutoff, two groups were differentiated. There were 1,028 who had a total score under six points (lower chronic emotional stress) and 1,145 with a total score equal to or over six points (higher chronic emotional stress). The group with higher chronic emotional stress was significantly younger (65 vs. 66 years, *p-value*<0.001), including significantly more females (66% vs. 57%, *p-value*<0.001) and black participants (14% vs. 11%, *p-value*=0.039). Participants with higher chronic emotional stress levels tend to have more depression (44% vs. 23%, *p-value*<0.001), and anxiety (24% vs. 10%, *p-value*<0.001). On a cardiometabolic level, more cases of T2DM (27% vs. 23%, *p-value*=0.028), CVD (8.7% vs. 6.4%, *p-value*=0.043), and obesity (50% vs. 42%, *p-value*<0.001) were recorded in the higher chronic stress group.

More tobacco (7.2% vs. 4.7%, *p-value*=0.014) and alcohol (1.1% vs. 0.3%, *p-value*=0.022) consumption were recorded in the higher chronic stress group, where a higher number of participants denied any form of regular physical activity (9.3% vs. 5.5%, *p-value*=0.001) and had higher IL-6 levels (−0.01 vs. −0.11 log-transformed value pg/mL, *p-value*=0.002). Further details are summarized in **Table 1**.

The comparison between different ethnic groups showed significantly higher chronic stress levels in Hispanics and Black participants compared to the White ones (**Figure 1.c**).

There was a significant positive log-shaped association between chronic stress and odds of depression and anxiety (*p* _non-linear models_ < 0.001) (**Figure 1.d and 1.e**).

### b. Chronic emotional stress and immunological and cardiometabolic biomarkers

Higher scores in chronic stress tests were significantly associated with an increase in BMI, HbA1c, and IL-6 levels, as well as higher values of systolic and diastolic blood pressure and cardiac rate. The associations remained statistically significant after adjusting for age, sex, and ethnicity. **Figure 2** shows that the effects might differ across different ethnic groups.

**Figure 2:**
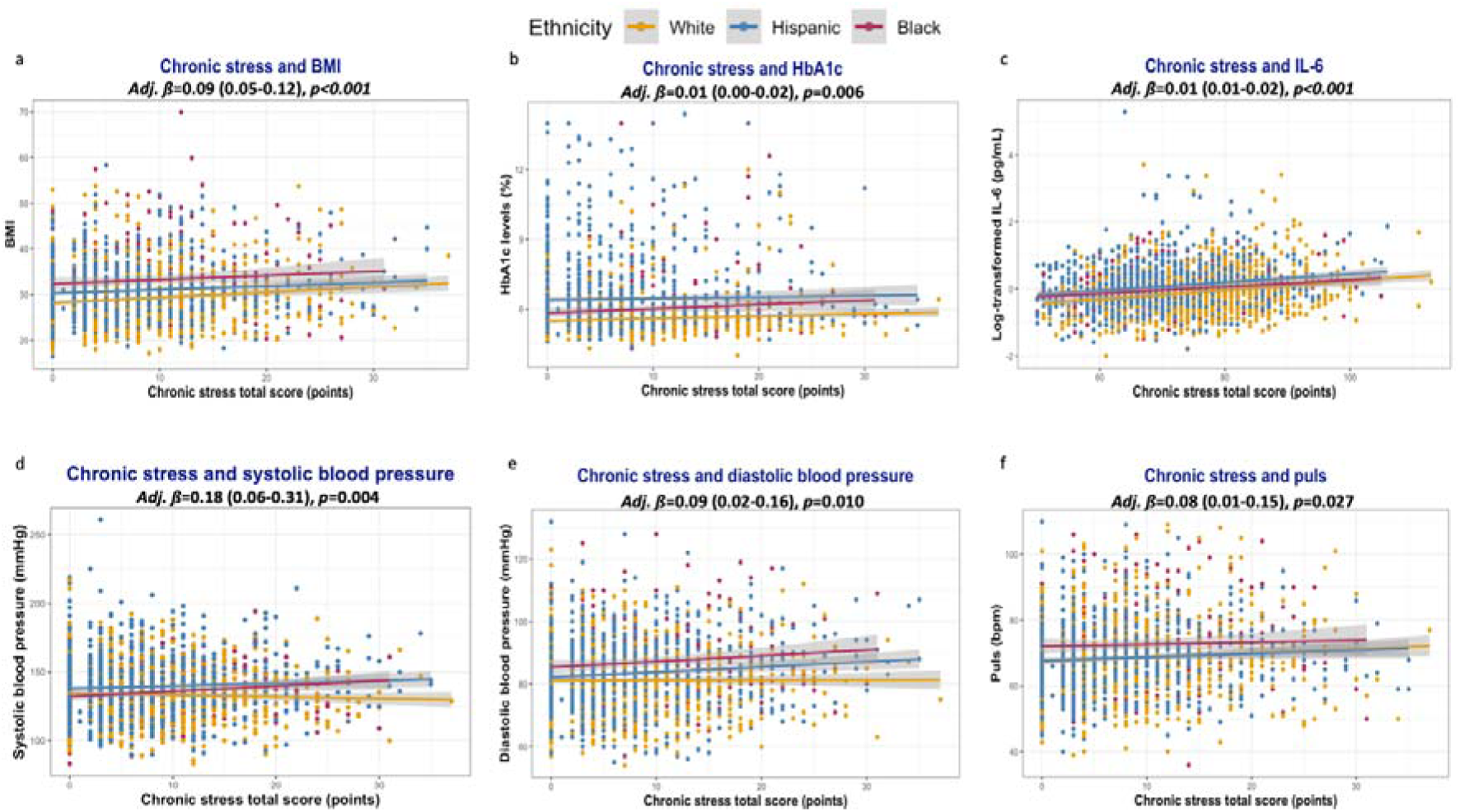
Associations between chronic emotional stress and biomarkers of cardiometabolic disorders. **2.a.** Body-mass index. **2.b.** HbA1c. **2.c.** Systemic Interleukine-6 levels. **2.d.** Systolic blood pressure. **2.e.** Systolic blood pressure. **2.f.** Heart rate.

### c. Emotional stress and cardiometabolic risks

Having a chronic stress total score equal to or over six points is associated with 53% higher odds of having concomitant CVD (adj. OR=1.53 [1.1-2.53]), 31% of T2DM (adj. OR=1.31 [1.06-1.62]), 23% of hypertension (adj. OR=1.23 [1.02-1.49]), and 30% obesity (adj. OR=1.3[1.09-1.55]). No significant association was found with dyslipidemia (**Figure 3.a**).

**Figure 3:**
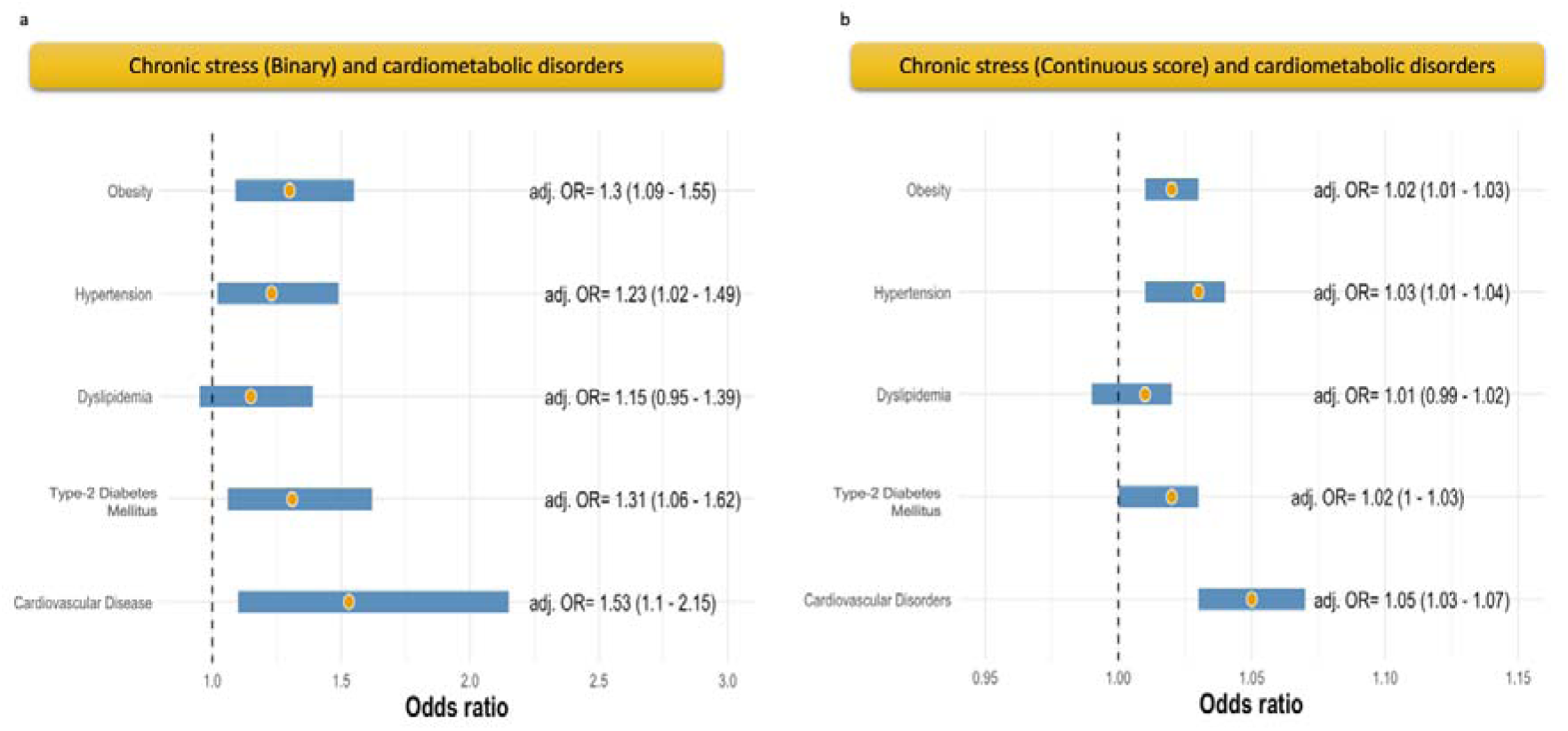
Associations between chronic emotional stress and different cardiometabolic disorders. **3.a.** Main analysis. **3.b.** Sensitivity analysis.

In the sensitivity analysis, the increase of one point in the total score of chronic emotional stress was linearly associated with a rise of 5% in the odds of CVD (adj. OR=1.05 [1.03-1.07]), 2% of diabetes (adj. OR=1.02 [1.00-1.03]), 3% of hypertension (adj. OR=1.03 [1.01-1.04]), and 2% of (adj. OR=1.02 [1.01-1.03]), with no significant association with dyslipidemia (**Figure 3.b**).

### d. Assessing mediating paths between chronic stress, IL-6, and cardiometabolic disorders

IL-6 mediated significantly 12% (*p-value*_FDR_=0.012) of the association between higher levels of chronic emotional stress and CVD, 17% with diabetes (*p-value*_FDR_<0.001), 18% with hypertension (*p-value*_FDR_<0.001), and 29% with obesity (*p-value*_FDR_=0.005) (**Figure 4.a**).

**Figure 4:**
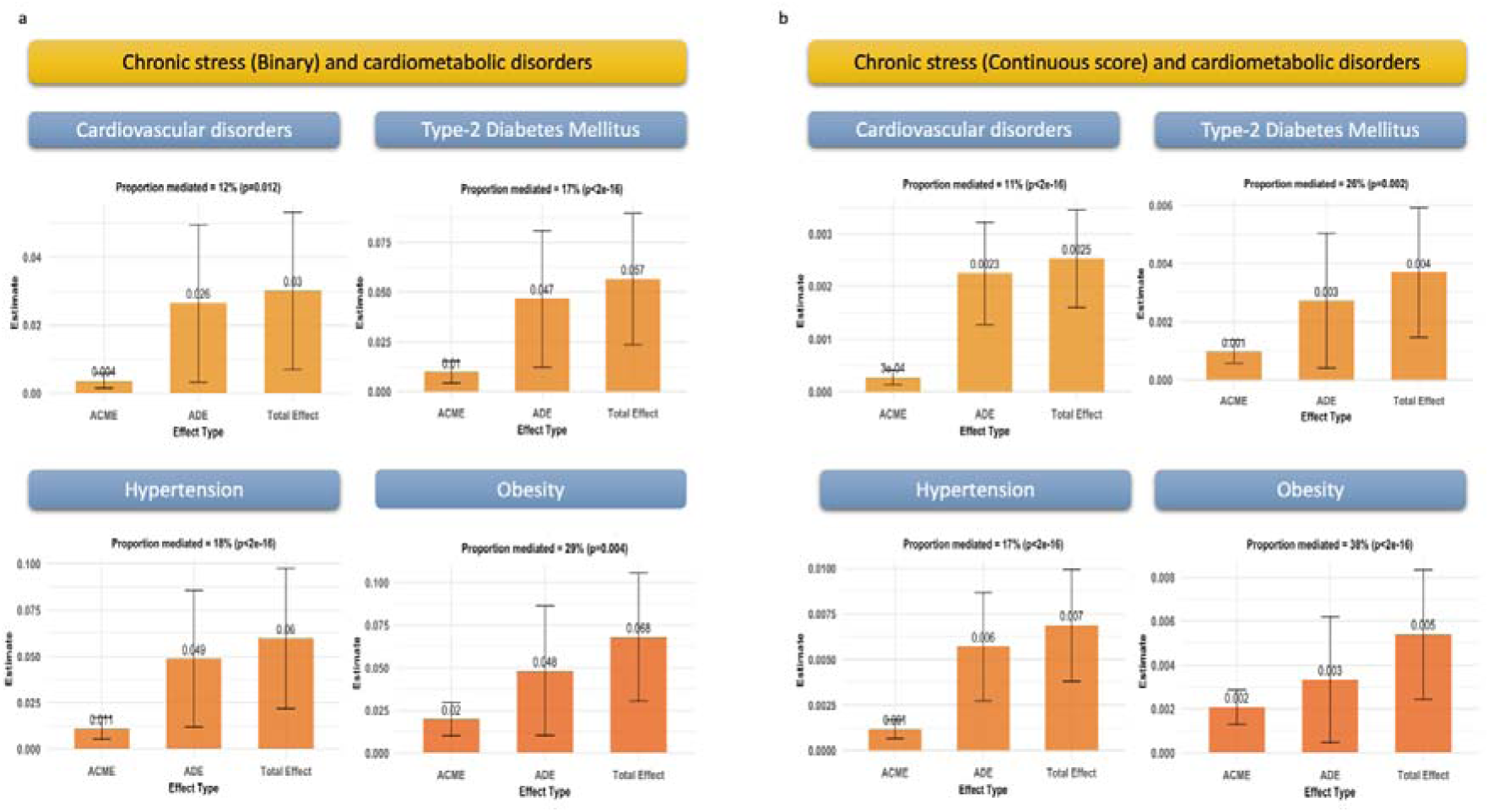
Mediation analysis of the effect of Interleukine-6 in the association between chronic emotional stress and different cardiometabolic disorders. **4.a.** Main analysis. **4.b.** Sensitivity analysis.

The sensitivity analysis using the continuous scores of chronic emotional stress as exposure showed comparable results (**Figure 4.b**).

## 4. Discussion

The study showed that chronic emotional stress had a strong predictive value on disclosing concomitant cardiometabolic disorders, mainly CVD, T2DM, hypertension, and obesity. These associations were partly mediated by IL-6, highlighting, statistically and non-exclusively, the value of low-grade systemic inflammation in the psychological-physical association.

### 4.1. Chronic emotional stress and cardiovascular risk

#### Chronic emotional stress

Chronic emotional stress can be triggered by several socioeconomic, behavioral, and physiological factors. (33, 34) Women engaged in shift work are more exposed to work-related stress and, consequently, to obesity. (34) An overwhelming, long-lasting working condition is associated with higher risks of several chronic diseases, such as diabetes, infections, and cardiovascular complications, in addition to an increased associated mortality risk. (35) Higher stress levels in people with chronic and severe diseases, such as breast cancer, are associated with higher depression rates and elevated systemic inflammation biomarkers, as well. (33)

The COVID-19 pandemic is the most recent example of a global health crisis impacting different health determinants. (36) Social and healthcare restrictions amplified the psychological burden of the crisis, and vulnerable populations, such as ethnic minorities, chronically ill persons, children, and women, were particularly affected. (37, 38) The COVID-19-related pandemic had a significant impact on the eating behavior and BMI of those exposed to increased stress. (39–43) Furthermore, natural disaster- or armed conflict-related traumatic stress is associated with a higher incidence of non-communicable physical disorders. (44)

#### Obesity

Childhood adversities, traumatic life events, and life stress in early adulthood are significantly associated with higher BMI. (45–47) Similarly, food insecurity-related stress is associated with higher BMI in adolescents and young adults. (48) Emotional eating, as a coping mechanism for facing stress, is a major cause of increased BMI, (49) also in black women. (50) The mediating role of pro-inflammatory cytokines was, however, rarely studied. In pregnant women, preexisting obesity was associated with higher systemic inflammatory biomarkers and perinatal depression risk. (51) Furthermore, pre-pregnancy BMI might play a mediating role between low socioeconomic status and higher IL-6 levels. (52) Thus, there is a serious lack of data on aging populations. The current analysis showed a significant association between chronic stress, higher IL-6 levels, and obesity.

#### Type-2 Diabetes Mellitus

The association between the onset and progression of T2DM and emotional stress is well recognized. (53) The association might be physiological through disturbing hormonal, inflammatory, and glucose homeostasis, or behavioral by negatively impacting health behaviors, mainly physical activity and nutrition. (53) Lower stress resilience at an early age is significantly associated with a higher risk of developing T2DM (HR=1.51), independently of BMI, family history, and socioeconomic risk factors. (54) In middle-aged adults, a low-variety diet associated with high emotional stress predicts higher odds of T2DM (OR=1.83 in men and 1.85 in females). (55) Psychological interventions in Latinos with T2DM showed an association between stress scores and HbA1c levels, which might reflect the importance of long-term and regular psychological support in stabilizing diabetes biomarkers and preventing long-term complications. (56)

#### Arterial hypertension

Increased sympathetic activity is a well-described mechanism through which emotional stress causes hypertension and heart rate variability. (57) Orexin might be a further mechanism involved in this association. (58) Emotional stress, depression, anxiety, insomnia, and hypertension might interact and impact the quality of life of affected persons. (59) In African Americans, emotional stress was significantly associated with a 15% to 22% higher risk of developing hypertension, independently from depression and anxiety. (60) In those with hypertension, emotional stress and depression were significantly associated with higher cardiometabolic factors. (61)

In the current analysis, depression and anxiety were not adjusted for in the model in order to estimate the overall effect of chronic emotional stress. Depression and anxiety are “intermediates” (or mediators) and not confounders in the path between emotional stress and cardiometabolic disorders. Adjusting for at least one of them reduces the results to a partial effect and underestimates global effects and eventual interactions.

#### Cardiovascular diseases

A study on a multiethnic population showed that work-related stress was a significant predictor of unfavorable cardiovascular health. (62) In multiethnic middle-aged and older adults, high psychological stress was significantly associated with cardiovascular disease, higher BMI, and depression. (63) Those factors, in addition to unfavorable health behaviors, significantly mediate the association between psychological stress and higher mortality. (63) In patients with stable coronary artery disease, even moderate emotional stress is associated with higher mortality. (64) The effect of stress starts early in life, and higher psychological stress in midlife is associated with a longitudinal increase in subclinical atherosclerosis during the follow-up. (65)

The association between stress, depression, anxiety, and cardiovascular disorders is also mediated by health behavior and nutrition. (66) A healthy lifestyle, including low-stress burdens and absence of depression, was significantly associated with a lower risk of atherosclerotic cardiovascular disease in a multiethnic population. (67) Psychotherapeutic interventions in patients with heart disease have beneficial effects on their mental and physical well-being, in addition to improving their quality of life. (68)

#### Depression and anxiety

Depression is significantly associated with cardiometabolic risks in different populations. (69–71) Similarly, lower depression frequencies were associated with a lower risk of cardiometabolic disease. (72) On the other side, several biomarkers of cardiometabolic risk showed significant associations with depression. (73–75) A meta-analysis showed that adults with a history of childhood maltreatment are three times more likely to develop depression or cardiometabolic disorders. (76) Preventive measures have demonstrated efficacy when the intervention was multidisciplinary and impacted physical and psychiatric risk factors. (77, 78)

### 4.2. Role of ethnicity

Belonging to a minoritarian ethnic group in any society is recognized to be an additional risk factor for health adversities, either owing to structural discrimination in health care coverage, higher exposure to risk factors, or a different physiological and genetic predisposition. (27, 29, 30) Comparing different patients with anxiety, non-Hispanic black ones had higher odds of being diagnosed with metabolic syndrome complications than non-Hispanic white ones. (79) Furthermore, older women with higher perceived discrimination express higher inflammation biomarkers, mainly IL-6, and higher IL-6 levels were significantly associated with higher BMI. (80)

### 4.3. Mediating role of low-grade inflammation

A very limited number of studies explored the association between chronic stress, systemic pro-inflammatory cytokines, and cardiometabolic disorders. (51, 52, 80) None of them was dedicated to older adults. This research gap highlights the importance of the current study in seeding awareness and motivating further research.

### 4.4. Strengths

The study included a high number of participants with complete data on relevant variables. Moreover, the diverse ethnic background of participants presents a major strength. By assessing the role of chronic stress, the study aims to highlight the importance of the psychosocial support of older populations, particularly members of socially marginalized groups. While this study focused on the role of chronic stress in predicting concomitant cardiometabolic diseases, which might be neglected and not investigated in medical examinations, it highlighted the statistical mediating effect of systemic inflammation in this association. The use of a non-parametric bootstrapping of 95% CI following the percentile method is a further methodological strength of the current results.

### 4.5. Limitations

The cross-sectional design is a major limitation of this study and does not allow for assessing causal effects. Chronic emotional stress was retrospectively reported by patients, which exposes the data to recall bias, and the exact duration of the stressful situation cannot be evaluated. Furthermore, the results have to be treated with precaution since the studied cardiometabolic disorders and IL-6 levels were only reported at baseline, and no priorly documented information on their past evolution is available. Most cardiometabolic disorders were diagnosed based on self-disclosure, and no medical records were revised in this community-based cohort. While causal relationship assumptions cannot be fulfilled with certainty in the current study owing to the unspecified chronological classification of events, the use of mediation analysis enables a better statistical understanding of the variations and interactions between variables. The study aimed to provide a preliminary theoretical framework for further large longitudinal clinic-based studies.

## 5. Conclusions

Depression and anxiety are far beyond being the unique adversities to which older persons belonging to ethnic minorities are exposed, and chronic emotional stress has to be explored in risk groups. The association of these psychological burdens with cardiometabolic disorders needs to be prioritized in a multidisciplinary and culturally sensitive medical approach. This implies that mental health management needs to be integrated into the preventive and curative strategies of physical healthcare. Psychological burdens have to be acknowledged in people suffering from cardiometabolic disorders, and psychotherapeutic support needs to be part of long-term secondary and tertiary prevention programs. While chronic emotional stress might play a relevant role in the onset and chronification of risks, mediating inflammatory factors need to be explored and assessed in causal frameworks to direct individualized therapeutic options.

## Declarations

### Ethical approval

All procedures contributing to this work comply with the ethical standards of the relevant national and institutional committees on human experimentation and with the Helsinki Declaration of 1975, as revised in 2013. Ethical approval was obtained from the local institutional reviewing board. Participants gave written informed consent. The current research is based on a secondary analysis of anonymized data and was performed in accordance with the data use agreement.

### Authorization for publication

The principal investigator and data administrator of the HABS-HD study reviewed the manuscript for its compliance with DUA and authorized the submission and publication of the current version.

### Authorship

AH has full access to all of the data and takes responsibility for the integrity of the data and the accuracy of the analysis, visualization, drafting, and editing of the manuscript.

### Data availability

Data can be acquired by qualified researchers after an official request.

## Acknowledgment

“Research reported on this publication was supported by the National Institute on Aging of the National Institutes of Health under Award Numbers R01AG054073, R01AG058533, R01AG070862, P41EB015922 and U19AG078109. The content is solely the responsibility of the authors and does not necessarily represent the official views of the National Institutes of Health.”

